# Effectiveness and Safety of Bempedoic Acid in Taiwanese Patients with Hypercholesterolemia ― A Pragmatic Phase IV Study (CLEAR Taiwan Study)

**DOI:** 10.64898/2026.02.06.26345788

**Authors:** I-Chang Hsieh, Dong-Yi Chen, Chih-Sheng Chu, Yi-Yao Chang, Bing-Hsiean Tzeng, Tien-Chi Huang, Heng-Hsu Lin, Wen-Po Chuang, Chi-Cheng Huang, Jih-Kai Yeh, Chun-Yuan Chu, Ming-Yun Ho, Ching-Ya Huang, Yu-Wen Chen, Wei-Chen Yang, Tsung-Hsien Lin, Yen-Wen Wu

**Author notes:** Correspondence: Yen-Wen Wu, Address: No. 21, Section 2, Nanya S. Road, Banqiao District, New Taipei City, Taiwan, Tsung-Hsien Lin, Address: No.100, Tzyou 1st Rd., Sanmin Dist., Kaohsiung City, Taiwan. These authors contributed equally to this work (Tsung-Hsien Lin, Yen-Wen Wu).

## Abstract

**Background:** Elevated low-density lipoprotein cholesterol (LDL-C) is a risk factor for cardiovascular disease. Despite available lipid-lowering therapies (LLT), lipid control remains suboptimal. Bempedoic acid offers a non-statin oral treatment for hypercholesterolemia. However, real-world data in Asia are limited. The study aimed to investigate the effectiveness and safety of bempedoic acid in Taiwan.

**Methods:** This pragmatic phase IV study enrolled 180 patients with inadequately controlled hypercholesterolemia to receive bempedoic acid for 12 weeks in addition to background LLT. The primary endpoint was the percentage change in LDL-C. Secondary endpoints included changes in other lipid parameters, high-sensitivity C-reactive protein (hsCRP), and safety outcomes.

**Results:** Among 180 patients, 160 (88.9%) completed the study. The median percentage change in LDL-C from baseline to week 12 was -19% (interquartile range [IQR]: -36.4% to -3.6%), decreasing from 117.5 to 92 mg/dL (*p* < 0.01). The median percentage changes from baseline to week 12 were -13.3% for non-high-density lipoprotein cholesterol (non-HDL-C), -10.8% for total cholesterol, -11.5% for apolipoprotein B, and -34.0% for hsCRP (all *p* < 0.01). Minimal effects were noted on triglycerides (+0.2%), HDL-C (-5.5%), and lipoprotein(a) (+2.6%) (all *p* > 0.05). At week 12, 31.3% of patients achieved LDL-C targets (< 100 mg/dL for primary prevention; < 55 or < 70 mg/dL for secondary prevention). The safety outcomes were consistent with the locally approved label, with no new safety signals identified.

**Conclusions:** Bempedoic acid offers an effective and safe oral therapeutic option for Taiwanese patients whose LDL-C levels remain inadequately controlled with existing LLT, including statins.

**Registration:** URL: https://clinicaltrials.gov/study/NCT06925100; Unique identifier: NCT06925100

**Clinical Perspective:** *What Is New?:*

◆ This pragmatic phase IV study provides the first real-world evidence from Taiwan demonstrating that bempedoic acid leads to clinically meaningful reductions in LDL-C (median percentage change: -19%) at week 12 when added to background lipid-lowering therapy in patients with inadequately controlled hypercholesterolemia.
◆ Approximately one-third of patients achieved guideline-recommended LDL-C targets within 12 weeks, with a safety profile consistent with the locally approved label and no new safety signals identified.

*What Are the Clinical Implications?:*

◆ Bempedoic acid represents an effective and well-tolerated oral add-on lipid-lowering option for Taiwanese patients who fail to achieve LDL-C goals with existing therapies, including those unable to tolerate or intensify statin treatment.

## INTRODUCTION

Cardiovascular disease (CVD) is one of the leading causes of death globally, responsible for nearly one-third of all deaths. Atherosclerotic cardiovascular disease (ASCVD), which accounts for approximately 85% of CVD cases, represents a major contributor to this global health burden [1]. In Taiwan, ASCVD is a significant public health concern, with an estimated 1.82 million people affected. Since 2014, the prevalence of ASCVD in Taiwan has increased steadily by 3.5%, a rate higher than that observed in other Asia-Pacific countries such as Japan (2.3%) and Australia (2.9%) [1]. Hyperlipidemia, the fastest-growing risk factor of ASCVD, affects 44% to 58% of Taiwanese adults [2].

Given the strong association between dyslipidemia and ASCVD, substantial reductions in cardiovascular risk can be achieved through effective management of modifiable risk factors—particularly low-density lipoprotein cholesterol (LDL-C). However, despite guideline recommendations, only 18.3% of high-risk primary prevention patients and 20.7% of secondary prevention patients achieved the treatment goals for LDL-C reduction based on the 2019 European Society of Cardiology (ESC) guidelines [3]. These real-world evidence point out the suboptimal implementation of ESC 2019 guidelines on LDL-C, in particular low use of combination lipid-lowering therapy (LLT), leading to a substantial proportion of patients remaining at high residual risk of ASCVD events [4].

Bempedoic acid, an oral, first-in-class novel non-statin lipid-lowering agent targeting LDL-C, received regulatory approval from the U.S. Food and Drug Administration and the European Medicines Agency in 2020 followed by approval from the Taiwan Food and Drug Administration in 2024. It is indicated for the treatment of primary hypercholesterolemia or mixed dyslipidemia as an adjunct to diet in combination with other LLTs. Bempedoic acid inhibits cholesterol biosynthesis through inhibition of adenosine triphosphate-citrate lyase, an upstream enzyme of 3-hydroxy-3-methylglutaryl-coenzyme A reductase in the cholesterol biosynthesis pathway. Like statins, bempedoic acid inhibits cholesterol synthesis and up-regulates LDL-C receptors. However, unlike statins, bempedoic acid is a prodrug and is not active in skeletal muscle tissue.

Randomized clinical trials have established the efficacy and acceptable safety profile of bempedoic acid, administered either as monotherapy or in combination with ezetimibe, yielding an average reduction in LDL-C ranging from 18% to 28% in patients with hypercholesterolemia [5]. Furthermore, the large CLEAR Outcomes trial, with 13,970 patients, demonstrated that bempedoic acid significantly reduced the risk of major adverse cardiovascular events by 13% compared to placebo among statin-intolerant patients [6].

However, existing clinical investigations of bempedoic acid have primarily concentrated on Western populations, with scarce data pertaining to East Asian cohorts and an absence of specific information regarding the Taiwanese population. To address this gap, the present phase IV interventional study aimed to evaluate the effectiveness and safety of bempedoic acid in Taiwanese patients, thereby providing insight into treatment outcomes within this demographic.

## METHODS

### Study Design

This was a pragmatic phase IV study carried out at three medical centers in Taiwan (NCT06925100). The study protocol included a screening phase lasting up to 14 days, followed by a 12-week open-label treatment period. Participants who discontinued the study were required to attend a follow-up visit within 2 to 4 weeks to facilitate continued safety assessment.

After obtaining written informed consent, investigators performed screening evaluations to assess patient eligibility. Enrolled patients received bempedoic acid 180 mg once daily for 12 weeks, prescribed at the discretion of the treating physician in routine clinical practice and in accordance with the locally approved label. During the study, patients were instructed to maintain their pre-existing LLT regimens to avoid confounding effects on laboratory parameters.

The study was conducted in accordance with the principles of the Declaration of Helsinki and the International Council for Harmonization’s Guideline for Good Clinical Practice. Approval was obtained from the ethics committees and institutional review boards of all participating institutions (Linkou Chang Gung Memorial Hospital: 202401853A4; Far Eastern Memorial Hospital: 113314-J; Kaohsiung Medical University Chung-Ho Memorial Hospital: KMUHIRB-F(1)-20240296). Informed consent was obtained from all participants prior to study enrollment.

### Participants

The study included adult patients aged 18 years and older who had been diagnosed with primary hypercholesterolemia (heterozygous familial or non-familial) or mixed dyslipidemia and who exhibited inadequately controlled LDL-C levels despite receiving LLT for a minimum of four weeks. Inadequate LDL-C control was defined as fasting LDL-C levels greater than or equal to 70 mg/dL for patients with ASCVD or those who had undergone coronary revascularization procedures, and greater than or equal to 100 mg/dL for patients outside of these categories. Patients were eligible if their treating physician intended to prescribe bempedoic acid according to the locally approved label, with no intervention from the study in the physician’s clinical judgment.

Patients with contraindications of bempedoic acid per locally approved label were not eligible for study participation, such as having estimated glomerular filtration rate (eGFR) less than 30 mL/min/1.73 m^2^, end-stage renal disease on dialysis, severe hepatic impairment (i.e., Child-Pugh C), or other contraindications to bempedoic acid (i.e., hypersensitivity to the active substance or any of the excipients, galactose intolerance, pregnancy, breastfeeding, or concurrent use of simvastatin exceeding 40 mg daily).

### Assessments and Endpoints

Clinical laboratory specimens for the assessment of fasting lipid profiles—including LDL-C, total cholesterol, high-density lipoprotein cholesterol (HDL-C), triglycerides, apolipoprotein B (apoB), and lipoprotein(a) [Lp(a)]—were obtained prior to dosing and at week 4 and 12 post-dosing. Samples for the measurement of glycosylated hemoglobin (HbA1c) and high-sensitivity C-reactive protein (hsCRP) were collected before dosing and at week 12 after doing. Safety evaluations included continuous monitoring of treatment-emergent adverse events (AEs), clinical laboratory parameters, vital signs, body weight changes, and physical examinations.

The primary effectiveness endpoint was the percentage change from baseline to week 12 in LDL-C levels. Secondary endpoints included percentage changes from baseline to week 12 in HDL-C, non-HDL-C, total cholesterol, triglycerides, apoB, and hsCRP, as well as the proportion of patients achieving LDL-C target levels at week 12. The LDL-C targets were defined as below 55 mg/dL and 70 mg/dL for patients with ASCVD or those who had undergone coronary revascularization procedures, in accordance with the 2022 Taiwan lipid guidelines for high-risk patients [7], and below 100 mg/dL for patients outside of these categories, based on the 2022 Taiwan lipid guidelines for primary prevention [8]. Safety endpoints comprised the incidence of AEs and adverse drug reactions related to bempedoic acid. Exploratory endpoints included the percentage change from baseline in Lp(a) and the change from baseline in HbA1c at week 12.

### Statistical Analysis

The study targeted a total of 180 patients. Phase III trial data demonstrated that bempedoic acid significantly reduced LDL-C by 19.2 to 21.8 mg/dL [9,10] in patients on maximally tolerated statin dose and 33.6 to 39.3 mg/dL [11,12] in statin-intolerant patients. A sample size of 44 patients was calculated to provide 80% statistical power to detect a significant LDL-C reduction of 19.2 mg/dL [9] at a type I error of 0.025 in statin-tolerant patients.

Similarly, a sample size of 16 patients was deemed sufficient to achieve 80% power to identify a significant LDL-C reduction of 33.6 mg/dL [11] at the same type I error in statin-intolerant patients. Given that approximately 10% of patients in Taiwan are statin-intolerant [13], the total sample size was set at 180 to ensure 160 completers, accounting for an anticipated dropout rate of 10%.

Descriptive statistical methods were applied to summarize the study results. Continuous variables were reported using number, mean, standard deviation (SD), median, interquartile range (IQR), range, and 95% confidence interval (CI). Categorical variables were described using counts and percentages. Changes from baseline were analyzed using either the paired-t test or the Wilcoxon signed rank test, as appropriate. No imputation methods were employed for missing data. Statistical significance was defined as p-value less than 0.05. All statistical analyses were performed using SAS^®^ (Statistical Analysis Software 9.4, SAS Institute Inc, Cary, North Carolina, USA).

## RESULTS

### Patient Characteristics

The study was conducted from April 2025 to October 2025 at three medical centers in Taiwan. A total of 180 patients were screened, and 180 eligible patients participated in the study. In all, 160 patients completed the study, with 20 patients discontinuing the study due to withdrawal of consent (14 patients) and AEs (6 patients). The mean (SD) adherence to study drug, as assessed by pill count, was 94.1% (14.6%).

The mean (SD) age was 61.4 (10.44) years and 62.7% of patients were male. Mixed dyslipidemia was identified in 76.7% of patients, 31.1% had diabetes, and 60.6% had hypertension. The mean (SD) baseline LDL-C was 118.1 (31.44) mg/dL and 67.2% of patients were secondary prevention patients who had pre-existing ASCVD.

Background statin therapy was used by 58.3% of patients (29.4% moderate-to-high intensity, 26.7% statin and non-statin LLT combinations), 22.2% were on background non-statin LLTs (the most common agent was ezetimibe) or very-low-to-low-dose statins, and 21.7% were not taking any LLTs. Nearly half of the population (46.1%) were classified as having statin intolerance (Table 1).

**Table 1.** Patient demographics and baseline characteristics.

| **Variables** | **All patients**<br>**(N = 180)** |
| --- | --- |
| Age, mean (SD), years | 61.4 (10.44) |
| < 65 years, n (%) | 112 (62.2) |
| ≥ 65 to < 75 years, n (%) | 54 (30.0) |
| ≥ 75 years, n (%) | 14 (7.8) |
| Male, n (%) | 113 (62.8) |
| BMI, mean (SD), kg/m<sup>2</sup> | 26.2 (4.35) |
| Medical history, n (%) |  |
| Coronary artery disease | 119 (66.1) |
| Hypertension | 109 (60.6) |
| Prior PCI | 75 (41.7) |
| Diabetes | 56 (31.1) |
| Heart failure | 34 (18.9) |
| Prior CABG | 21 (11.7) |
| Atrial fibrillation | 19 (10.6) |
| Chronic kidney disease* | 17 (9.4) |
| Liver disease<sup>†</sup> | 16 (8.9) |
| Prior MI | 16 (8.9) |
| Secondary prevention, n (%)<sup>‡</sup> | 121 (67.2) |
| Background LLTs, n (%)<sup>§</sup> |  |
| All statin use | 105 (58.3) |
| Statin monotherapy | 57 (31.7) |
| Statin alone (high intensity) | 14 (7.8) |
| Statin alone (moderate intensity) | 39 (21.7) |
| Statin alone (very-low-to-low intensity) | 4 (2.2) |
| Statin combinations | 48 (26.7) |
| Statin + ezetimibe | 39 (21.7) |
| Other statin combinations | 9 (5.0) |
| All non-statin use | 36 (20.0) |
| Ezetimibe alone | 21 (11.7) |
| Other non-statin LLT combinations | 15 (8.3) |
| PCSK9i combinations | 0 (0.0) |
| No LLT documented | 39 (21.7) |
| Statin intolerant, n (%) | 83 (46.1) |
| Baseline lipid levels, median (IQR), mg/dL |  |
| LDL-C | 117.5 (92.5, 137.5) |
| Total cholesterol | 182.5 (158.0, 206.5) |
| non-HDL-C | 133.5 (107.0, 157.5) |
| HDL-C | 47.0 (39.0, 58.0) |
| Triglyceride | 119.0 (84.0, 164.5) |
| Apolipoprotein B | 95.0 (79.0, 111.0) |
| Lipoprotein(a) | 13.9 (5.3, 40.0) |
| Baseline hsCRP, median (IQR), mg/L | 0.9 (0.4, 1.8) |

\* Chronic kidney disease (CKD) was defined as having diagnosis of CKD or estimated glomerular filtration rate < 60 mL/min/1.73 m<sup>2</sup>.

| Variables | All patients<br>(N = 180) |
| --- | --- |

<sup>†</sup> Liver diseases included hepatitis, hepatic impairment, and cirrhosis.
<sup>‡</sup> Secondary prevention was defined as patients who had pre-existing atherosclerotic cardiovascular disease (coronary heart disease, stroke, and/or peripheral arterial disease)
<sup>§</sup> Statin intensity classification is based on the 2013 American College of Cardiology/American Heart Association guidelines [27].
<sup>||</sup> Statin intolerance was defined as using low-dose or very-low-dose statin with or without other concurrent LLT, stopping all statin use, or using ezetimibe monotherapy or other non-statin monotherapy, either continued or ceased, irrespective of prior statin use, at the time of screening.
Abbreviations: BMI, body mass index; SD, standard deviation; PCI, percutaneous coronary intervention; CABG, coronary artery bypass graft; MI, myocardial infarction; LLT, lipid-lowering therapy; IQR, interquartile range; LDL-C, low-density lipoprotein cholesterol; HDL-C, high-density lipoprotein cholesterol; non-HDL-C, non-high-density lipoprotein cholesterol; PCSK9i, proprotein convertase subtilisin/kexin type 9 inhibitor

### Effectiveness

Bempedoic acid significantly reduced LDL-C levels (*p* < 0.01). The median (IQR) percentage change in LDL-C from baseline to week 12 was -19% (-36.4% to -3.6%), decreasing from a median (IQR) of 117.5 mg/dL (92.5 to 137.5 mg/dL) to 92 mg/dL (72 to 115 mg/dL) (Figure 1A). Over 12 weeks of treatment, a divergence in LDL-C response was observed (Figure 2). For patients with an LDL-C increase of > 10%, poor adherence (treatment compliance < 60%) accounted for 25% of cases. In addition, as adherence to background LLT was not systematically monitored, inconsistent use of concomitant LLT, including missed doses or intentional discontinuation due to pill burden, may partly explain the paradoxical LDL-C increases observed in these patients.

**Figure 1.**
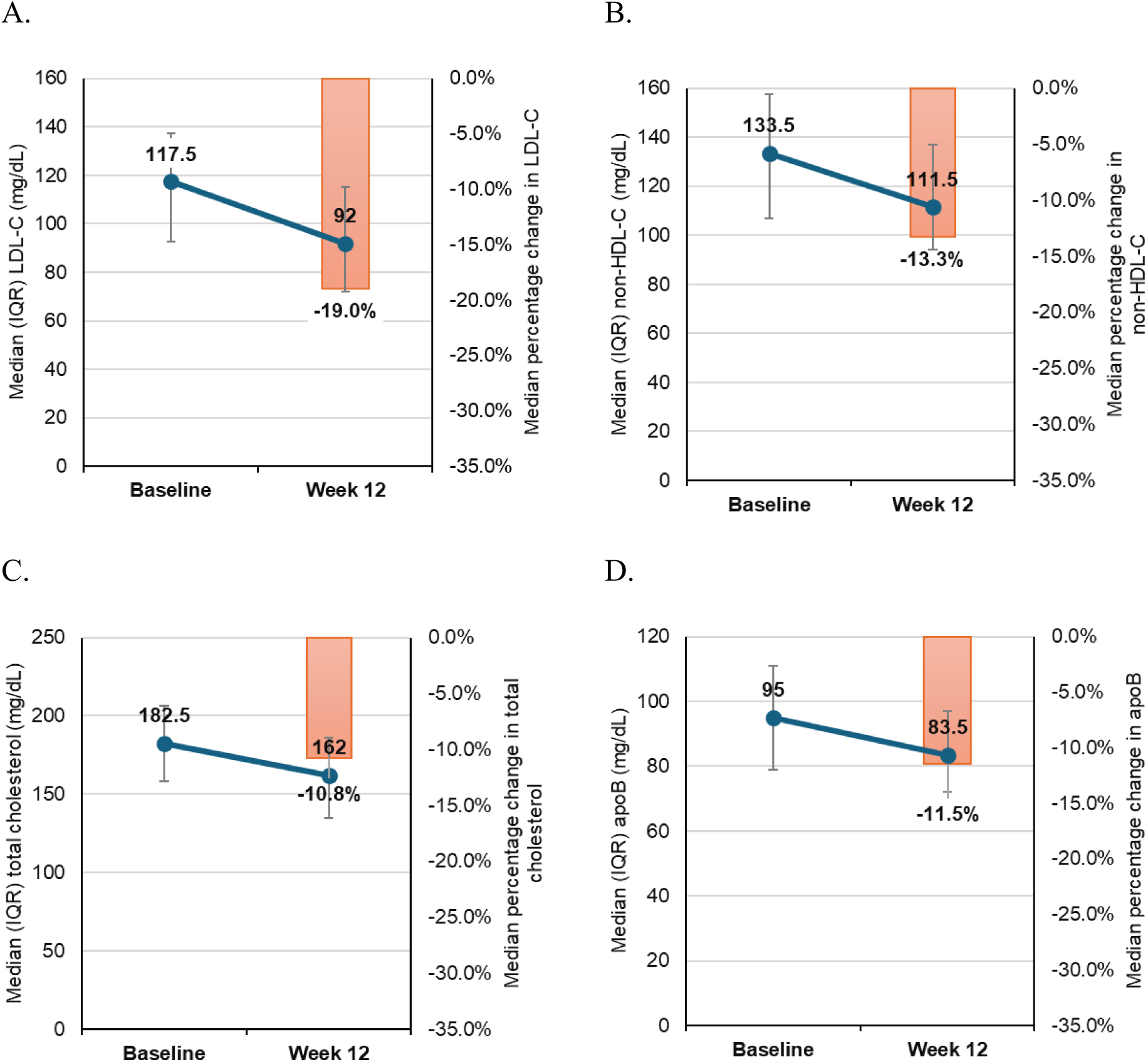
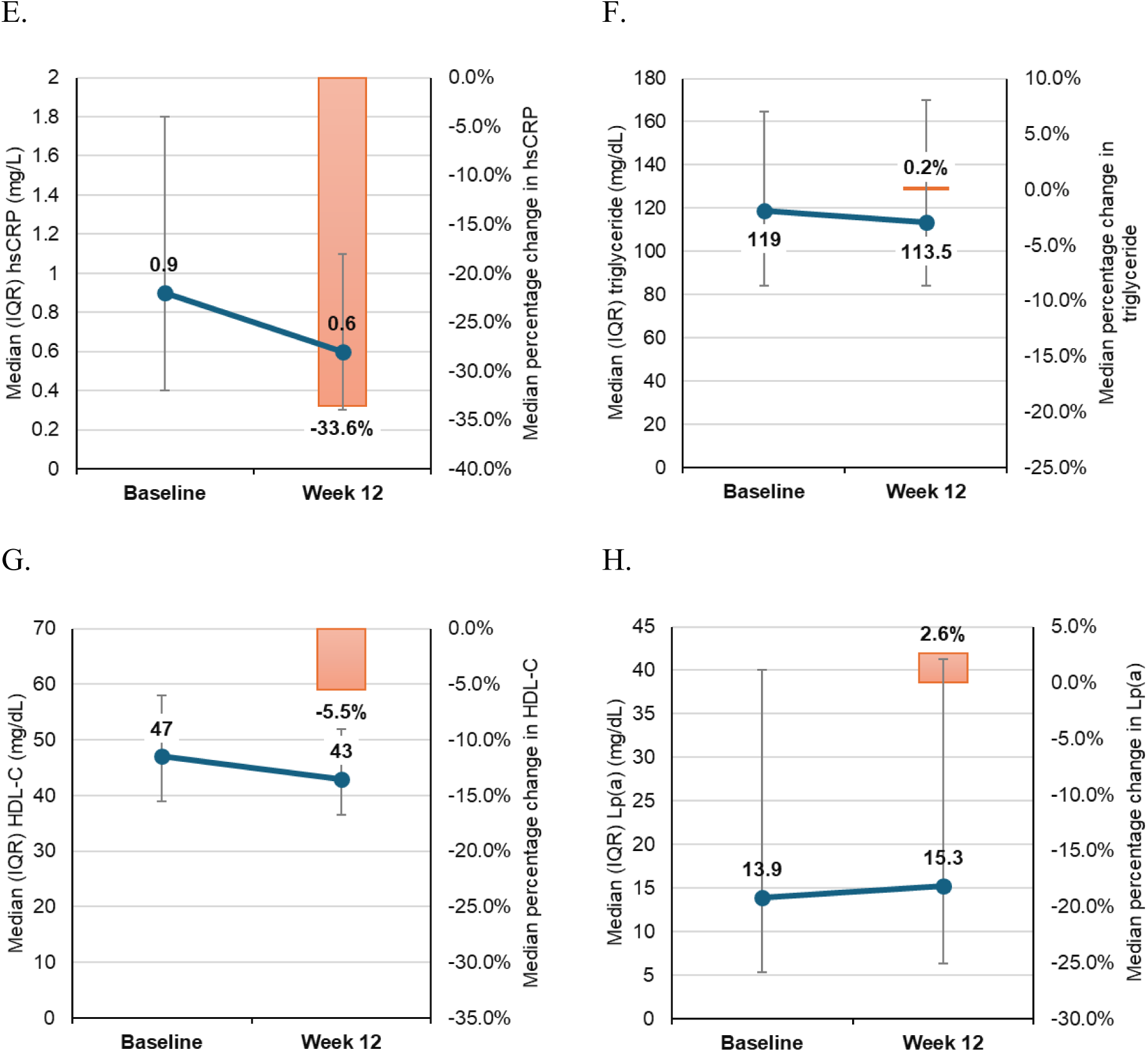
Levels and percentage change from baseline to week 12 in lipid parameters and hsCRP. Data are presented as median with interquartile range (IQR). Blue dots and lines are median levels of laboratory tests, while orange bars represent the percentage change from baseline. A. low-density lipoprotein cholesterol (LDL-C); B. non-high-density lipoprotein cholesterol (non-HDL-C); C. total cholesterol; D. apolipoprotein B (apoB); E. high-sensitivity C-reactive protein (hsCRP); F. triglycerides; G. high-density lipoprotein cholesterol (HDL-C); H. lipoprotein(a) (Lp(a))

**Figure 2.**
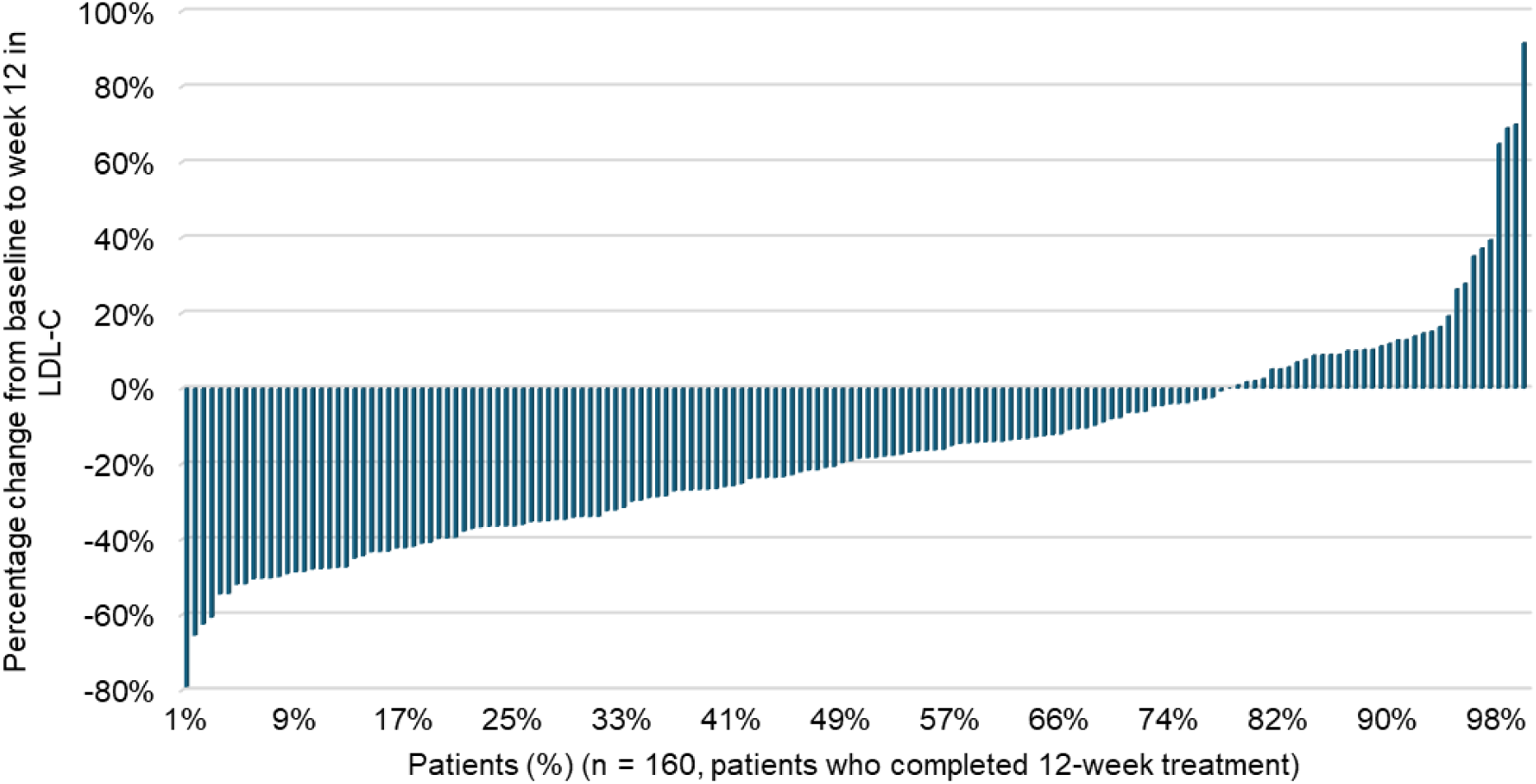
Waterfall plot showing distribution of percentage change in low-density lipoprotein cholesterol (LDL-C) levels at week 12. Patients are ordered from highest to lowest LDL-C reduction along the x-axis, which represents the cumulative percentage of patients.

Significant reductions were also observed for other lipid parameters at week 12 (*p* < 0.01). The median (IQR) percentage changes from baseline to week 12 were -13.3% (-26.8% to 4.5%) for non-HDL-C, -10.8% (-22.2% to 1.8%) for total cholesterol, and -11.5% (-23.1% to 2.4%) for apoB (Figure 1B, C, D). Marked reductions from baseline in hsCRP were also observed at week 12 (*p* < 0.01), with a median (IQR) change of -33.6% (-54.7% to 0%) (Figure 1E). Effects on triglycerides, HDL-C, and Lp(a) were minimal (median change from baseline at week 12: 0.2%, -5.5%, 2.6%, respectively; Figure 1F, G, H).

At baseline, 28.9% of patients achieved an LDL-C level below 100 mg/dL, while none achieved the targets of less than 70 mg/dL or less than 55 mg/dL. By week 12, these percentages rose to 59.4%, 19.4%, and 9.4%, respectively (Figure 3). Among high-risk patients with pre-existing ASCVD, the percentages attaining LDL-C levels below 70 mg/dL and below 55 mg/dL were 24.3% and 11.2%, respectively, at week 12 (Figure 3).

**Figure 3.**
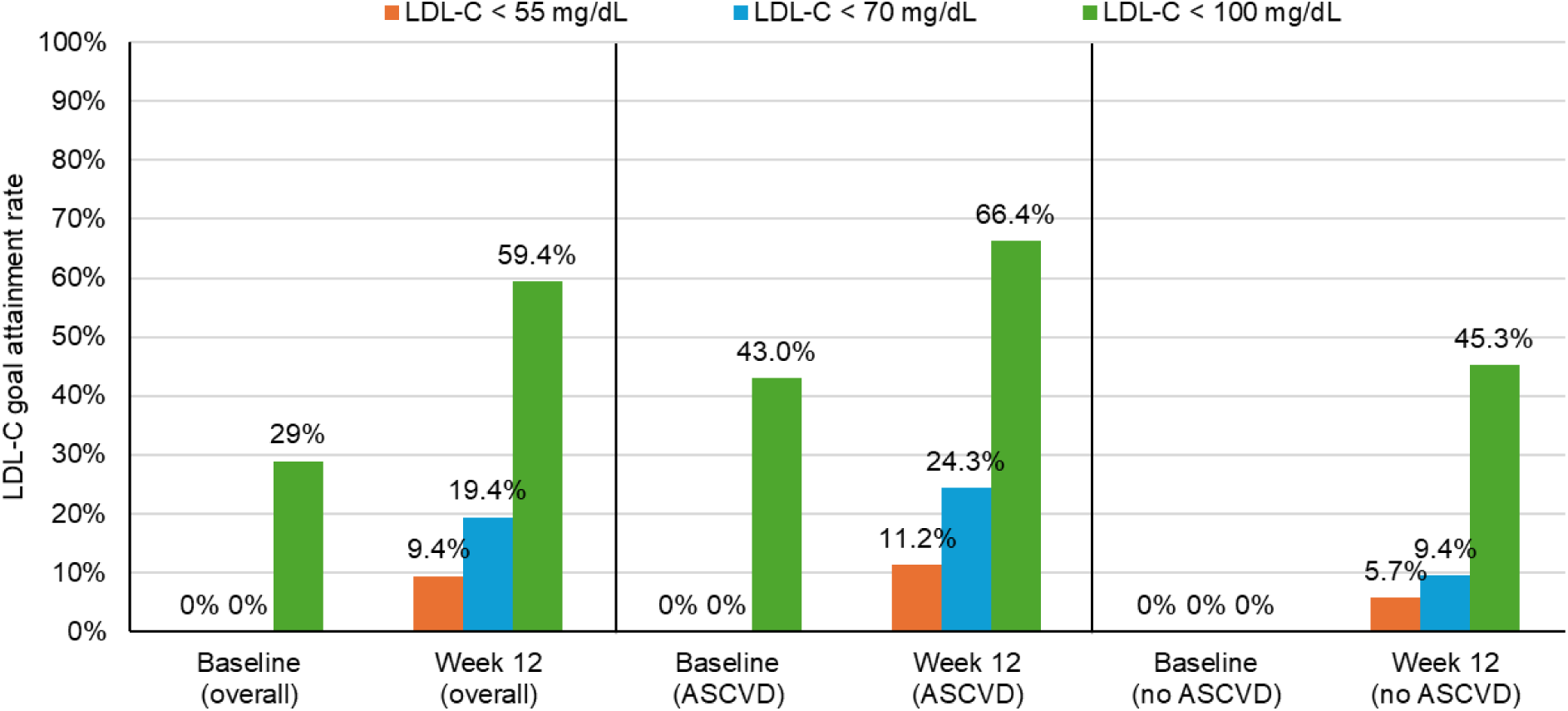
Low-density lipoprotein cholesterol (LDL-C) goal attainment rate at baseline and week 12 for total population (n = 180), patients with pre-existing atherosclerotic cardiovascular disease (ASCVD), defined as coronary heart disease, stroke, and/or peripheral arterial disease (n = 121), and patients without pre-existing ASCVD (n = 59).

### Safety

A total of 85 AEs occurred in 35% of patients. Almost all AEs were mild (84 out of 85), with no serios AEs reported across the 12-week study period and 56.5% of AEs (48 out of 85) were judged to be unrelated to the study drug (Table 2).

**Table 2.** Adverse events.

| Variables | Event count | All patients<br>(N = 180), n<br>(%) |
| --- | --- | --- |
| Any AEs | 85 | 63 (35.0) |
| Mild | 84 | 62 (34.4) |
| Moderate | 1 | 1 (0.6) |
| Serious AEs | 0 | 0 (0.0) |
| Study drug-related AEs | 37 | 34 (18.9) |
| Discontinuation of study drug | 6 | 6 (3.3) |
| Most common AEs* |  |  |
| AST or ALT clinically significant increase<sup>†</sup> | 18 | 16 (8.9) |
| - ALT or AST > 3 × ULN | 2 | 2 (1.1) |
| Uric acid clinically significant increase<sup>†</sup> | 7 | 7 (3.9) |
| - Increase in uric acid > 2.0 mg/dL | 3 | 3 (1.7) |
| Triglycerides clinically significant increase<sup>†</sup> | 6 | 6 (3.3) |
| - Triglycerides > 2 × baseline value | 2 | 2 (1.1) |
| Arthralgia | 5 | 5 (2.8) |
| Weight decreased (decrease from baseline > 5%) | 4 | 4 (2.2) |
| Laboratory results |  |  |
| AST, U/L, mean (SD) change, baseline to week 12 |  | 7.4 (15.22) |
| ALT, U/L, mean (SD) change, baseline to week 12 |  | 4.7 (22.36) |
| Uric acid, mg/dL, mean (SD) change, baseline to week 12 |  | 1.00 (1.221) |
| Creatinine, mg/dL, mean (SD) change, baseline to week 12 |  | 0.080.207) |

\* Occurred in ≥ 2% of patients
<sup>†</sup> Judged as clinically significant increase by physicians
Abbreviations: AE, adverse events; ULN, upper limit of normal; AST, aspartate aminotransferase; ALT, alanine aminotransferase; SD, standard deviation

AEs leading to treatment discontinuation occurred in 6 (3.3%) patients, including elevated liver enzyme (n = 2), skin rashes (n = 1), increased triglycerides (n = 1), hyperuricemia (n = 2), and gastrointestinal fullness (n = 1). Except for gastrointestinal fullness, these AEs were considered by investigators to be related to the study drug, but not by patients’ symptoms or specific laboratory values.

AEs observed in ≥ 2% of patients included increased aspartate aminotransferase (AST) or alanine aminotransferase (ALT) (8.9% [n = 16]), increased uric acid (3.9% [n = 7]), increased triglycerides (3.3% [n = 6]), arthralgia (2.8% [n = 5]), and decreased body weight (2.2% [n = 4], mean percentage change: -6.7%); all of which were asymptomatic. Marked elevations in AST or ALT (> 3 × upper limit of normal [ULN]), triglycerides (> 2 × baseline), and uric acid (change from baseline > 2.0 mg/dL) were observed in two (1.1%), three (1.7%), and two (1.1%) patients, respectively (Table 2). Myalgia was reported in three patients (1.7%), and all cases were considered mild and not clinically significant (data not shown).

The mean change from baseline to week 12 was 7.4 U/L for AST, 4.7 U/L for ALT, 1.0 mg/dL for uric acid, and 0.08 mg/dL for creatinine (Table 2). No patients reported gout, experienced increases in total bilirubin > 2 × ULN, or met Hy’s law criteria. No clinically meaningful changes were observed in other laboratory parameters, vital signs, or physical examination findings.

## DISCUSSION

To our knowledge, this is the first phase IV study to describe the use of bempedoic acid in Asian patients with inadequately controlled primary hypercholesterolemia or mixed dyslipidemia. In Taiwanese patients receiving background lipid-lowering therapy, the addition of bempedoic acid resulted in a clinically meaningful 19% reduction in LDL-C from baseline to week 12, which is comparable to reductions reported in phase III pivotal trials conducted predominantly in Western populations (-20.3% [95% CI: -23.5 to -17.1]) [14].

Additionally, treatment with bempedoic acid enabled 31.1% of patients to achieve guideline-recommended LDL-C targets, including LDL-C < 100 mg/dL in 45.3% of patients without prior ASCVD and LDL-C < 70 mg/dL in 24.3% of those with prior ASCVD. Bempedoic acid was generally well tolerated, with most patients experiencing no adverse events and demonstrating high treatment adherence. The safety profile observed in this study was consistent with previous clinical trials and the locally approved label, with no new safety signals identified.

Our findings extend the existing evidence base by demonstrating similar LDL-C-lowering efficacy in an Asian population. In randomized phase III trials of patients with ASCVD and/or heterozygous familial hypercholesterolemia, bempedoic acid achieved mean placebo-corrected LDL-C reductions of -17.8% in statin-treated patients and -24.5% in statin-intolerant patients after 12 weeks of treatment [15]. Similarly, in the CLEAR-J trial, which evaluated bempedoic acid over 12 weeks in Japanese patients, LDL-C levels were reduced by 21.8% [16]. In the present pragmatic phase IV study, a comparable LDL-C-lowering effect was observed at week 12, with a median reduction of -19% in the overall population, -18.5% in statin-tolerant patients, and -22.3% in statin-intolerant patients (data not shown). Beyond randomized trials, our findings are also consistent with results from other phase IV and real-world observational studies. In the European observational MILOS study, 524 German patients treated with bempedoic acid or bempedoic acid/ezetimibe fixed-dose combination (FDC) for one year achieved a mean LDL-C reduction of 27.3%, with the proportion of patients reaching LDL-C goals increasing from 4.0% before treatment to 25.8% at one year [17]. Similarly, in a Belgian cohort of the MILOS study comprising 375 patients, bempedoic acid or bempedoic acid/ezetimibe FDC treatment resulted in a mean LDL-C reduction of 22.7% at week 8, and the LDL-C goal attainment rate increased from 5.0% at baseline to 33.7% at week 8 [18]. Although the present study did not include patients receiving bempedoic acid/ezetimibe FDC, the magnitude of LDL-C lowering and the improvement in goal attainment observed in our cohort were broadly comparable, with the proportion of patients achieving guideline-recommended LDL-C targets increasing to 31.1% at week 12. Collectively, the concordance between our results and those of prior randomized and observational studies supports the clinical utility and generalizability of the LDL-C-lowering effects of bempedoic acid across diverse patient populations.

In the present study, substantial inter-individual variability in LDL-C reduction was observed with bempedoic acid treatment. This heterogeneity may be attributable to genetic variations in the *ACLY* gene, which encodes ATP citrate lyase, differences in drug activation mediated by very-long-chain acyl-CoA synthetase 1 (ACSVL1), and inter-individual variability in the pharmacokinetics of bempedoic acid. Furthermore, polymorphisms in genes regulating cholesterol homeostasis may also contribute to the observed variability. Genetic determinants of lipid-lowering response have been well documented for statins; for example, polymorphisms in the apolipoprotein E gene have been associated with variation in statin-induced LDL-C reduction [19], and variants in transporter genes such as *SLCO1B1* have been linked to differential lipid responses to statin therapy [20]. Given that bempedoic acid and statins target different enzymes within the same cholesterol biosynthesis pathway, it is plausible that similar genetic factors may also modulate the lipid-lowering efficacy of bempedoic acid. Regarding non-biological factors, despite overall treatment adherence was high in the current study (mean adherence, 94%), the study did not collect data regarding diet, lifestyle modifications, or adherence to other background LLT. Suboptimal use of background LLT, such as missed doses or reluctance to take multiple medications, may have contributed to the observed LDL-C fluctuations. These findings highlight the need for future studies to identify the determinants of response to bempedoic acid and to optimize personalized lipid-lowering strategies in Asian populations.

The effects of bempedoic acid on other lipid parameters observed in the present study were generally consistent with findings from prior clinical investigations. Meta-analyses of randomized pivotal studies have demonstrated that bempedoic acid is associated with significant reductions in total cholesterol (-12.4%), apolipoprotein B (-13.2% to -14.3%), and non-HDL-C (-15.3% to -15.5%), while exhibiting minimal effects on triglycerides, HDL-C, and Lp(a) [14,21,22].

Regarding inflammatory biomarkers, the hsCRP reduction observed in the current study (-33.6%) appears to be numerically greater than that reported in most of the prior studies (approximately -23.4% to -31.9%) [14,16,11]. This finding may indicate a potentially augmented anti-inflammatory effect within the current study population. Mechanistically, preclinical research has revealed that bempedoic acid mediates anti-inflammatory effects by activating AMP-activated protein kinase (AMPK) in primary human monocyte-derived macrophages and in in vivo inflammation models via a liver kinase B1 (LKB1)-dependent pathway [23]. Despite these anti-inflammatory effects have not yet been fully established in humans, the greater reduction in hsCRP observed in the Taiwanese cohort relative to Western populations may be attributable to multiple factors, including variations in baseline inflammatory status, genetic background, dietary patterns, and cardiometabolic risk profiles. In addition, emerging data suggest that genetic polymorphisms affecting inflammatory signalling and cholesterol metabolism may modulate the pleiotropic effects of statins [24].

Given that both bempedoic acid and statins target the same cholesterol biosynthesis pathway, population-specific genetic differences may also contribute to the observed heterogeneity in inflammatory responses. Nonetheless, owing to the pragmatic, real-world nature of the current study and its relatively short follow-up period, these hypotheses remain speculative and necessitate further investigation through pharmacogenomic and mechanistic studies.

Our study underscores a significant unmet medical need among patients with hypercholesterolemia in Taiwan. By design, the study exclusively enrolled patients whose LDL-C levels remained inadequately controlled despite ongoing lipid-lowering therapy. Notably, although nearly 70% of the cohort consisted of high-risk patients undergoing secondary prevention, only 29.4% were treated with moderate- to high-intensity statin monotherapy, and 35.0% received combination LLTs. Furthermore, approximately half of the patients met the study-defined criteria for statin intolerance, suggesting a susceptibility to intolerance of other LLTs, potentially influenced by the nocebo effect [25], which may contribute to suboptimal LDL-C management. Within this context, bempedoic acid provides a well-tolerated and effective oral therapeutic alternative, addressing this unmet need through an additional mechanism to further reduce LDL-C levels in patients who fail to achieve guideline-recommended targets with current LLTs.

The effectiveness of bempedoic acid observed in the current study was accompanied by a favourable safety profile. No serious adverse events were reported, and occurrences of muscular disorders were rare (1.7%, n = 3) and uniformly mild. This favorable muscle safety profile is biologically plausible, given that bempedoic acid functions as a prodrug activated by ACSVL1, an enzyme predominantly expressed in the liver but absent in skeletal muscle [26]. Accordingly, bempedoic acid constitutes a suitable LLT option for patients who experience muscle-related intolerance to statins.

Despite asymptomatic elevations in hepatic enzymes and uric acid levels were noted, the incidence of AST or ALT elevations > 3 × ULN (1.2%) and increased uric acid (3.9%) were lower than those reported in a prior 12-week phase III trial (3.9% and 7.7%, respectively) [11]. Previous studies have indicated that bempedoic acid is linked to modest elevations in serum uric acid levels. This effect is attributed to the weak inhibition of bempedoic acid and its active metabolite on the renal organic anion transporter 2, which plays a role in uric acid excretion [26]. From a clinical perspective, caution may therefore be warranted when prescribing bempedoic acid to patients with a history of gout or hyperuricemia. Nonetheless, despite the relatively short 12-week observation period and the lack of long-term data in the Taiwanese cohort, the uric acid elevations observed in the current study did not raise major clinical concerns, and no cases of gout were reported. While the CLEAR Outcomes trial documented slight elevations in the occurrence of cholelithiasis and tendon rupture [6], no such adverse events were observed among patients in the current study. Taken together, these safety findings support the clinical utility of bempedoic acid as a well-tolerated add-on LLT within Asian clinical settings.

The strengths of the current study lie in its distinction as the first phase IV evaluation of bempedoic acid within an Asian cohort and the only data in Taiwanese patients. The study provides relevant information on the initial clinical adoption of the new oral LLT, bempedoic acid, among Asian patients. By adopting a pragmatic study design, applying eligibility criteria aligned with the locally approved label, and closely mirroring the clinical practice, this study enhances its relevance and generalizability to routine clinical settings in Taiwan. This investigation offers valuable insights into the application of this novel non-statin lipid-lowering agent among patients exhibiting suboptimal LDL-C control despite ongoing LLT and requiring additional interventions to achieve optimal cardiovascular risk mitigation.

However, certain limitations should be acknowledged, including the nonrandomized design, absence of a comparator arm, and the relatively short follow-up period. Therefore, further large-scale and extended observational studies are necessary to comprehensively assess the long-term effects of bempedoic acid on lipid parameters, cardiovascular outcomes, and safety profiles in Asian populations in real-world settings.

## CONCLUSION

Bempedoic acid represents an effective and well-tolerated oral treatment option for Taiwanese patients with inadequately controlled LDL-C levels. Importantly, these findings provide valuable clinical evidence to support the use of bempedoic acid in lipid management among Asian populations.

## Data Availability

Data collected within this study will be made available to researchers after contacting the corresponding author and upon approval by the study sponsor. The researcher should describe the required data and purposes in a proposal. A data sharing agreement will be set up by the study sponsor. All data provided are anonymized to respect the privacy of patients who have participated in the trial in line with applicable laws and regulations.

## ACKNOWLEDGMENTS

The authors would like to express their appreciation to all sub-investigators and participating site study teams for their collaboration and dedication to this multicenter study.

## SOURCES OF FUNDING

This work was supported by Daiichi Sankyo Taiwan, Ltd.

## DISCLOSURES

The authors have read and confirmed their agreement with the International Committee of Medical Journal Editors authorship and conflict of interest criteria. The authors have also confirmed that this article is unique and not under consideration by nor published in any other publication and that they have permission from rights holders to reproduce any copyrighted material. CY Huang, YW Chen, and WC Yang are employees of Daiichi Sankyo Taiwan Ltd. All other authors have declared no conflicts of interest.

## AUTHOR CONTRIBUTIONS

IC Hsieh, YW Wu, TH Lin, CY Huang, YW Chen, and WC Yang developed the study concept and design. IC Hsieh, YW Wu, TH Lin, DY Chen, CS Chu, YY Chang, BH Tzeng, TC Huang, HH Lin, WP Chuang, CC Huang, JK Yeh, CY Chu, and MY Ho acquired the data. IC Hsieh, YW Wu, TH Lin, CY Huang, and YW Chen analyzed the data. CY Huang wrote the first draft of the manuscript. IC Hsieh, YW Wu, and TH Lin provided critical revision of the manuscript for important intellectual content. DY Chen, CS Chu, YY Chang, BH Tzeng, TC Huang, HH Lin, WP Chuang, CC Huang, JK Yeh, CY Chu, MY Ho, YW Chen, and WC Yang jointly developed the structure and arguments of the paper. All authors reviewed and approved the final manuscript.

## Non-standard Abbreviations and Acronyms

LLT: lipid-lowering therapy
ACSVL1: very-long-chain acyl-CoA synthetase 1
FDC: fixed-dose combination
AMPK: AMP-activated protein kinase
LKB1: liver kinase B1

## Notes

### Clinical Trial

ClinicalTrials.gov. ID: NCT06925100

### Clinical Protocols

https://clinicaltrials.gov/study/NCT06925100

### Author Declarations

Participating centers and the IRB approval number: Linkou Chang Gung Memorial Hospital: 202401853A4 Far Eastern Memorial Hospital: 113314-J Kaohsiung Medical University Chung-Ho Memorial Hospital: KMUHIRB-F(1)-20240296

